# Circulating amino acids, amino acid metabolites, dipeptides, and other cationic metabolites and risk of breast cancer

**DOI:** 10.1101/2020.09.01.20185835

**Authors:** Oana A. Zeleznik, Raji Balasubramanian, Yibai Zhao, Lisa Frueh, Sarah Jeanfavre, Julian Avila-Pacheco, Clary B. Clish, Shelley S. Tworoger, A. Heather Eliassen

## Abstract

**Background:** Breast cancer is the most common malignancy among women in the United States, with more than 250,000 cases diagnosed each year. Metabolomics, which reflect the aggregate effects of genetics and the environment on an individual’s metabolic state, can shed light on biochemical pathways involved in susceptibility to breast cancer. We investigated associations between pre-diagnostic circulating amino acids-related metabolites and subsequent risk of breast cancer among predominantly premenopausal women.

**Methods:** In 1996-1999, 29,611 women (average age, 44 years) in the Nurses’ Health Study II donated blood samples. Between blood collection and June 2011, 1057 women were diagnosed with breast cancer (average of 8 years after blood collection). Women were predominately premenopausal at the time of blood collection. 207 amino acid and amino acid-related metabolites were profiled with LC-MS/MS. Conditional logistic regression (CLR) was used to estimate odds ratios (ORs) of breast cancer and 95% confidence intervals (CIs). Multivariable analyses evaluating the joint association of all metabolites with breast cancer risk were based on CLR with a lasso penalty (Lasso), CLR with an elastic net penalty (Elastic Net), and Random Forests. We used FDR to account for testing multiple hypotheses.

**Results:** Eleven metabolites were associated with breast cancer risk in CLR models, after adjustment for multiple comparisons (p value < 0.05 and q value < 0.20; creatine had q value > 0.20), 6 of which remained significant after adjustment for breast cancer risk factors (p-value<0.05). Higher levels of six metabolites, including 2-aminohippuric acid, DMGV, kynurenic acid, N2, N2-dimethylguanosine, phenylacetyl glutamine and piperine, were associated with lower breast cancer risk (e.g., piperine: OR_simple_ (95%CI) = 0.85 (0.78-0.93); OR_adjusted_ (95%CI)=0.84 (0.77-0.92)). Higher levels of asparagine, creatine and 3 lipids (C20:1 LPC, C34:3 PC plasmalogen, C40:7 PE plasmalogen) were associated with increased breast cancer risk (e.g., C40:7 PE plasmalogen OR_simple_ (95%CI) = 1.14 (1.05-1.25); OR_adjusted_ (95%CI) = 1.11 (1.01-1.22)). Piperine, 2-aminohippuric acid, C40:7 PE plasmalogen and creatine were also selected in multivariable modeling approaches (Lasso, Elastic Net, and Random Forests).

**Conclusions:** Two diet-related metabolites, piperine (responsible for the pungency of pepper) and 2-aminohippuric acid (the glycine conjugate of the tryptophan metabolite anthranilic acid) were inversely associated, while C40:7 PE plasmalogen (a highly unsaturated glycerophospholipid and key component of the lipid bilayer of cells) was positively associated with breast cancer risk among predominately premenopausal women, independent of established breast cancer risk factors. Further validation of the specific metabolite associations with breast cancer risk in independent cohorts is warranted.

## Introduction

Breast cancer is the most common malignancy among women in the United States, with more than 250,000 cases diagnosed each year^1^. Known modifiable risk factors are estimated to account for only around one-third of postmenopausal breast cancers^2-4^, and an even smaller fraction of premenopausal cancers^2,5^. Thus, new strategies are needed for the identification of modifiable risk factors, especially for premenopausal breast cancers.

Metabolites are small molecules that are produced and consumed by cellular metabolism. The study of the complete collection of metabolites, called metabolomics, provides a direct signature of cellular activity in the body and has emerged as a powerful tool for the diagnosis, characterization, and prediction of disease. Metabolomic methods have uncovered biomarkers for a wide variety of cancers including colorectal, gastric, pancreatic, liver, ovarian, breast, urinary, esophageal and lung^6^. In breast cancer, metabolomics has proven useful for tumor biology characterization, predicting treatment response, anticipating recurrence, and estimating prognosis ^7^.

More recently, prospective epidemiological studies have used metabolomics to identify metabolite risk factors for several cancers including pancreatic^8-10^, prostate^11,12^, liver^13^, colorectal^14^, ovarian^15,16^, endometrial^17^, and breast cancer^18-22^. For breast cancer, studies have used both targeted^18,21,23^ and untargeted^20,22^ methods to discover metabolomic risk factors associated with diet^20,23^, BMI^21^, microbiota metabolism^20^, lipid, amino acid, and other metabolic pathways^18,20,22^. While several studies stratified results by estrogen receptor (ER) status^18,21,23^, no prospective metabolomic breast cancer studies have investigated differential effects by menopausal status.

In this study, we assessed the association of over 200 prospectively measured circulating amino acid and amino acid-related metabolites with risk of breast cancer among the predominantly premenopausal women (1057 cases and 1057 matched controls) of the Nurses’ Health Study II (NHSII).

## Methods

### Study Population

In 1989, 116,429 female registered nurses aged 25-42y returned a mailed questionnaire and were enrolled in the NHSII. Participants have been followed biennially since 1989 with questionnaires collecting information on reproductive history, lifestyle factors, diet, medication use, and new disease diagnoses.

In 1996-1999, 29,611 NHSII participants aged 32-54y contributed blood samples, as previously described^24^. Of these, 18,521 women who had not used oral contraceptives, been pregnant or breastfed in the previous six months provided samples timed within the menstrual cycle, targeting the early follicular (days 3 to 5 of the cycle) and mid-luteal (7 to 9 days prior to expected start of next cycle) phases. The remaining women donated a single untimed sample. Follicular plasma was separated and frozen by the participants and returned with the luteal sample; samples were collected and shipped overnight to our laboratory where we processed and archived aliquots of white blood cell, red blood cell, and plasma in liquid nitrogen freezers (≤-130°C). Follow-up in the blood subcohort is high (96% in 2011).

The study protocol was approved by the institutional review boards of the Brigham and Women’s Hospital and Harvard T.H. Chan School of Public Health, and those of participating registries as required. The return of the self-administered questionnaire and blood sample was considered to imply consent.

### Case and Control Selection

Cases of breast cancer were identified after blood collection among women who had no reported cancer (other than nonmelanoma skin). 1057 cases (invasive cases n=780) were diagnosed between 1999 and 2011. Breast cancer cases were reported by the participant, which were confirmed by medical record reviews (n=1015) or verbally by the nurse (n=42). Given the high confirmation rate by medical record for breast cancer in this cohort (99%), all cases are included in this analysis.

One control was matched per case by the following factors: age (+/- 2y), menopausal status and postmenopausal hormone therapy (HT) use at blood collection and diagnosis (premenopausal, postmenopausal and not taking HT, postmenopausal and taking HT, and unknown), and month (+/- 1mo), time of day (+/- 2h), fasting status at blood collection (<8 h after a meal or unknown; >10h), race/ethnicity (African-American, Asian, Hispanic, Caucasian, other) and luteal day (+/- 1d; timed samples only).

### Covariate Information

Data on breast cancer risk factors, including anthropometric measures, reproductive history, and lifestyle factors, were collected from questionnaires administered biennially and at the time of blood collections. Case characteristics, including invasive vs. *in situ*, histologic grade, estrogen and progesterone receptor (ER, PR), were extracted from pathology reports. As previously described^25^, immunohistochemical results for ER and PR, read manually by a study pathologist, were included for cases with available tumor tissue included in tissue microarrays.

### Laboratory Assay

Plasma metabolites were profiled at the Broad Institute of MIT and Harvard (Cambridge, MA) using a liquid chromatography tandem mass spectrometry (LC-MS) method designed to measure polar metabolites such as amino acids, amino acids derivatives, dipeptides, and other cationic metabolites as described previously^26-29^ . Pooled plasma reference samples were included every 20 samples and results were standardized using the ratio of the value of the sample to the value of the nearest pooled reference multiplied by the median of all reference values for the metabolite. Samples were run together, with matched case-control pairs (as sets) distributed randomly within the batch, and the order of the case and controls within each pair randomly assigned. Therefore, the case and its control were always directly adjacent to each other in the analytic run, thereby limiting variability in platform performance across matched case-control pairs. In addition, 238 quality control (QC) samples, to which the laboratory was blinded, were also profiled. These were randomly distributed among the participants’ samples.

Hydrophilic interaction liquid chromatography (HILIC) analyses of water soluble metabolites in the positive ionization mode were conducted using an LC-MS system comprised of a Shimadzu Nexera X2 U-HPLC (Shimadzu Corp.; Marlborough, MA) coupled to a Q Exactive mass spectrometer (Thermo Fisher Scientific; Waltham, MA). Metabolites were extracted from plasma (10 µL) using 90 µL of acetonitrile/methanol/formic acid (74.9:24.9:0.2 v/v/v) containing stable isotope-labeled internal standards (valine-d8, Sigma-Aldrich; St. Louis, MO; and phenylalanine-d8, Cambridge Isotope Laboratories; Andover, MA). The samples were centrifuged (10 min, 9,000 x g, 4°C), and the supernatants were injected directly onto a 150 x 2 mm, 3 µm Atlantis HILIC column (Waters; Milford, MA). The column was eluted isocratically at a flow rate of 250 µL/min with 5% mobile phase A (10 mM ammonium formate and 0.1% formic acid in water) for 0.5 minute followed by a linear gradient to 40% mobile phase B (acetonitrile with 0.1% formic acid) over 10 minutes. MS analyses were carried out using electrospray ionization in the positive ion mode using full scan analysis over 70-800 m/z at 70,000 resolution and 3 Hz data acquisition rate. Other MS settings were: sheath gas 40, sweep gas 2, spray voltage 3.5 kV, capillary temperature 350°C, S-lens RF 40, heater temperature 300°C, microscans 1, automatic gain control target 1e6, and maximum ion time 250 ms. Metabolite identities were confirmed using authentic reference standards or reference samples.

In total, 259 known metabolites were measured in this study. Metabolites not passing our previously conducted processing delay pilot study ^29^ were excluded from this analysis (N=33). All metabolites (N=226) included here exhibited good reproducibility within person over 1-2 years^29^. 206 metabolites had no missing values among participant samples. One metabolite had <10% missing values and 19 metabolites had ≥10% missing values. Most of the metabolites (N=191) had a coefficient of variation (CV) <25% and an intraclass correlation coefficient (ICC) >0.4 among blinded QC samples. Twenty-five metabolites had CV≥25%, five had ICCs≤0.4, and five metabolites had CV≥25% and ICC≤0.4.

### Statistical Analysis

Metabolite levels were natural logarithm transformed and standardized prior to statistical analysis. Missing values were imputed by one half the lowest observed value per metabolite, for metabolites with <10% missing values (N=1). Metabolites with >10% missing values (N=19) were excluded from the main analysis and evaluated in an exploratory analysis.

Association of metabolite levels with breast cancer risk was assessed in metabolite-by-metabolite models and in multivariable analyses that included all metabolites simultaneously.

The association of individual metabolites with breast cancer risk was assessed in conditional logistic regression models. In a simple model, each metabolite was included without adjustment for other factors. In an adjusted model, the following additional factors were included: BMI at age 18, weight change from age 18 to time of blood draw, age at menarche, parity and age at first birth, family history of breast cancer, diagnosis of benign breast disease, physical activity, alcohol consumption, exogenous hormone use and breastfeeding history. Odds ratios (OR) and 95% confidence intervals (95% CI) were estimated for a one-unit (one standard deviation) increase in the log-transformed and standardized metabolites levels.

We performed analyses restricting to premenopausal women at blood collection, and analyses stratified by BMI (<25 vs. ≥25 kg/m^2^) and ER status. In a sensitivity analysis, we observed similar results between conditional logistic regression and unconditional logistic regression adjusting for the matching factors. Thus, stratified analyses were conducted using unconditional logistic regression, additionally adjusting for the matching factors. To test for effect modifications by BMI and ER status, we included cross-product terms in conditional logistic models and report the p-value for that interaction. As ER status represents a case characteristic, we assigned each control the ER status of its matched case.

In an exploratory analysis we assessed the association with risk of breast cancer for the 19 metabolites with >10% missing values. We included the continuous metabolite level as well as a presence/absence indicator in the fully adjusted conditional logistic regression model and performed a likelihood-ratio test (full model compared to a model excluding both the metabolite and the presence/absence indicator) to estimate the significance level of the association.

Multivariable analyses evaluating the joint association of the 207 metabolites with breast cancer risk were based on (1) conditional logistic regression with lasso penalty (‘Lasso’), (2) conditional logistic regression with an elastic net penalty (‘Elastic Net’), and (3) Random Forests. In the Lasso and Elastic Net analyses, a minimally adjusted model included only the set of 207 metabolites, whereas a fully adjusted model further adjusted for the risk factors noted above. In each analysis, the optimal values of the regularization parameter(s) were estimated as that which minimizes the average deviance in the left-out partitions, in a 10-fold cross validation procedure. A p-value for each metabolite was obtained from a permutation test in which the case/control labels were permuted within each matched stratum. A p-value for each metabolite was calculated as the proportion of permutations (out of 250) in which the magnitude of the coefficient under label permutation was at least as large as the regression coefficient in the observed dataset. Analyses were carried out using the R library clogitL1^30^.

Random Forests analyses included a minimally adjusted model that included the 207 metabolites and matching factors. A fully adjusted model also included the additional risk factors noted above. In all analyses, the parameter *mtry* corresponding to the number of variables randomly sampled as candidates at each split was set to the square root of the total number of covariates in the model. Each classifier was an aggregate of 5000 trees. A p-value for each metabolite was obtained from a permutation test as described above in which the case/control labels were randomly permuted 100 times. Analyses were carried out using the R library randomForest^31^.

To adjust for multiple testing in the conditional logistic regression and the multivariable models we estimated the positive FDR based on the q-value procedure^32^. Metabolites that satisfied a p-value less than 0.05 and corresponding q-value less than 0.20 in the minimally adjusted model were discussed as primary findings.

Criterion for statistical significance: Metabolites that met a p-value<0.05 and q-value<0.20 in at least one of the four models (Conditional Logistic Regression, Lasso, Elastic Net and Random Forests) with minimal adjustment were considered as statistically significant. An exception was made for metabolites that did not meet a p-value threshold < 0.05 in the conditional logistic regression: 4 metabolites that met the threshold for statistical significance in Lasso only but had high raw p-values (>0.3) in the conditional logistic regression were excluded (C12:1 carnitine, C22:5 LPC, C46:2 TAG, glycine).

A metabolite score was estimated for each participant as a linear combination of all metabolites that met the threshold for statistical significance. The coefficients associated with each metabolite were estimated in a conditional logistic regression model with the Lasso penalty that included all metabolites simultaneously and with full adjustment for all potential confounders.

## Results

### Study population

1057 cases and 1057 matched controls were included in this study (Table 1). Women were an average 53 years old and predominantly premenopausal (80%) at the time of blood collection. At diagnosis, 42% of the women were premenopausal and 46% were postmenopausal.

**Table 1:** Characteristics of breast cancer cases and matched controls at blood collection in the Nurses’ Health Study II, mean (SD) or %.

|  | **Cases**<br>**(n=1057)** | **Controls**<br>**(n=1057)** |
| --- | --- | --- |
| **Age at blood collection<sup>^</sup>, y** | 44.7 (4.5) | 44.8 (4.4) |
| **Age at menarche, y** | 12.4 (1.3) | 12.8 (1.4) |
| **Parity and age at first birth, %:** |  |  |
| Nulliparous | 21.1 | 15.9 |
| 1-2 children, $\geq 25$y | 39.2 | 34.9 |
| 1-2 children, $< 25$y | 14.7 | 15.9 |
| 3+ children, $< 25$y | 11.3 | 16.6 |
| 3+ children, $\geq 25$y | 13.8 | 14.2 |
| **Ever breastfed, %:** | 63.1 | 65.0 |
| **Family history of breast cancer, %:** | 17.4 | 10.8 |
| **Personal history of benign breast disease, %:** | 22.1 | 15.6 |
| **BMI at age 18, kg/m<sup>2</sup>** | 20.8 (2.9) | 21.1 (3.1) |
| **Weight change between age 18 and blood collection, kg** | 11.6 (12.0) | 12.6 (13.2) |
| **Physical activity, MET-hrs/wk** | 18.0 (15.3) | 18.1 (15.5) |
| **Alcohol consumption, g/day** | 3.8 (6.9) | 3.3 (5.6) |
| **Past/current exogenous hormone use\*, %:** | 86.3 | 86.7 |
| **Menopausal status at blood collection<sup>^</sup>, %:** |  |  |
| Premenopausal | 80.2 | 79.7 |
| Postmenopausal | 12.7 | 13.1 |
| Unknown | 7.1 | 7.3 |
| **Menopausal status at diagnosis<sup>^</sup>, %:** |  |  |
| Premenopausal | 42.0 | 42.2 |
| Postmenopausal | 46.4 | 47.1 |
| Unknown | 11.6 | 10.7 |
| **Caucasian<sup>^</sup>, %:** | 97.2 | 98.4 |
| **Fasting($> 8$h) at blood collection<sup>^</sup>, %** | 68.7 | 74.7 |

\* oral contraceptive or menopausal hormone therapy
<sup>^</sup> matching factor

### Conditional logistic regression (CLR)

Eleven metabolites were significantly associated with risk of breast cancer based on the simple model (Figure 1, Supplementary Table 1). Six metabolites were associated with lower risk while five metabolites were associated with higher risk of overall breast cancer. Dimethylguanidino valeric acid (DMGV; OR per 1-SD increase (95%CI)=0.84 (0.77-0.92)), 2-aminohippuric acid (OR (95%CI)=0.84 (0.76-0.92)) and piperine (OR (95%CI)=0.85 (0.78-0.93)) had the strongest inverse associations. C40:7 phosphatidylethanolamine (PE) plasmalogen (OR (95%CI)=1.14 (1.05-1.25)) and asparagine (OR (95%CI)=1.14 (1.04-1.26)) had the strongest positive associations. Creatine was the only metabolite with q-value>0.2 in the simple model but is included here as q-value<0.2 in both Lasso models. Results were similar when we included adjustment for breast cancer risk factors (DMGV: 0.88 (0.79-0.97); 2-aminohippuric acid: 0.85 (0.77-0.93); C40:7 PE: plasmalogen: 1.11 (1.01=1.22); asparagine: 1.10 (1.00-1.22)) and when we restricted to premenopausal women (Figure 1, Supplementary Table 2) or ER+ tumors (Figure 1, Supplementary Table 3).

**Figure 1:**
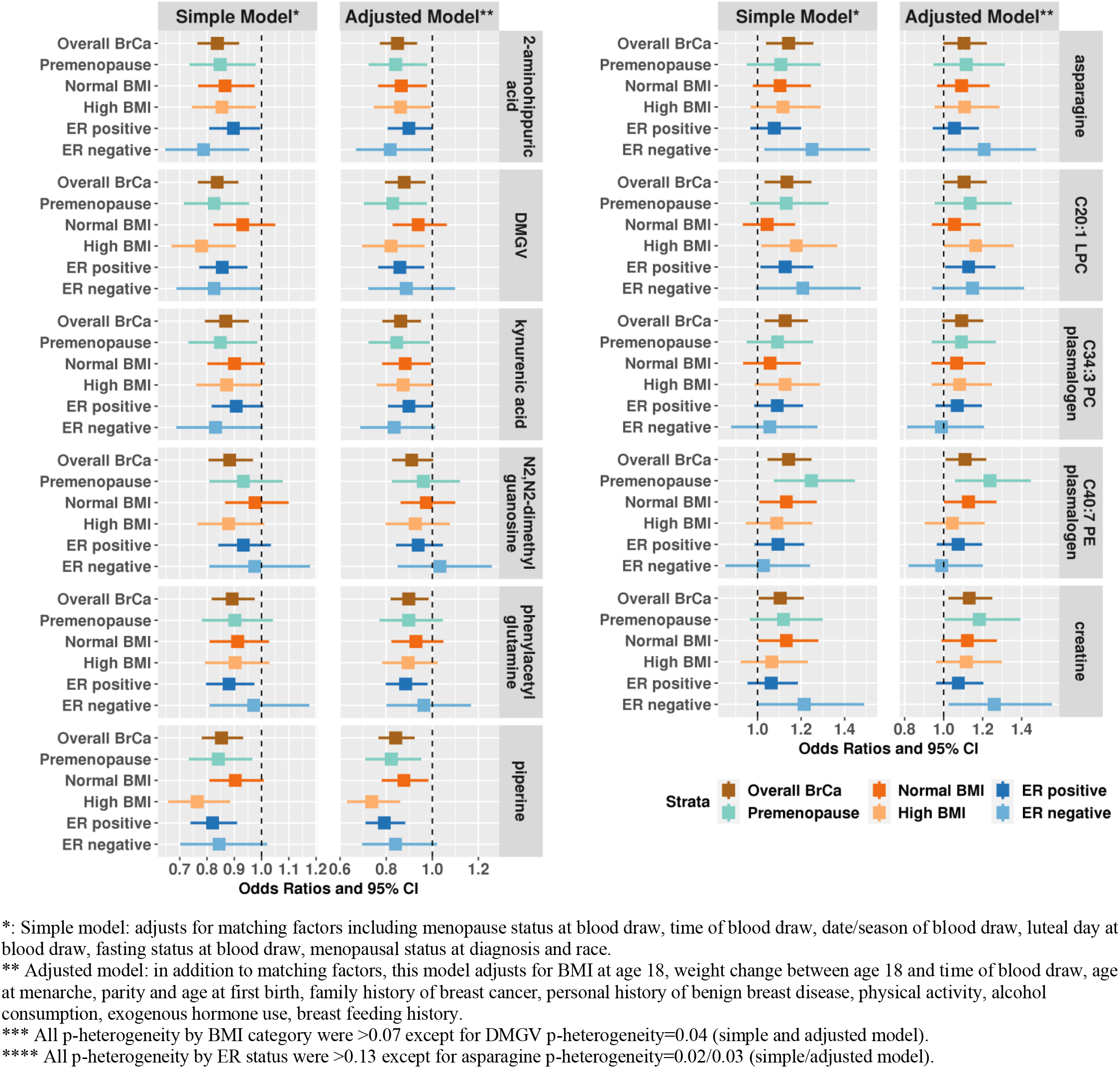
Odds ratios and 95% confidence intervals (CI) per 1 SD increase for metabolites significantly associated with risk of overall breast cancer (p-value<0.05 and q-value<0.2) in Nurses’ Health Study II, among premenopausal women only, by BMI category (<25, ≥25)^***^, and by ER status (ER positive, ER negative)^****^. Creatine, although not selected by the conditional logistic regression (p-value<0.05, q-value>0.2), is shown here for completeness, as is was selected by the multivariable models.

Among the 11 selected metabolites, only DMGV showed effect modification by BMI with stronger associations among women with high BMI (adjusted model, high BMI: OR (95%CI)=0.82 (0.70-0.97); normal BMI: OR (95%CI)=0.94 (0.83-1.06); p-interaction=0.04). Significant interactions with BMI were observed for several additional metabolites that were not significant overall; for example C36:4 DAG/TAG fragment (high BMI: OR (95%CI)=0.92 (0.80-1.06); normal BMI: OR (95%CI)=1.21 (1.06-1.37); p-interaction=0.005), serine (high BMI: OR (95%CI)=1.19 (1.04-1.38); normal BMI: OR (95%CI)=0.93 (0.83-1.05); p-interaction=0.008), and proline betaine (high BMI: OR (95%CI)=0.82 (0.72-0.94); normal BMI: OR (95%CI)=1.04 (0.92-1.17); p-interaction=0.017).

Of the 11 selected metabolites, we only observed stronger associations with risk of ER- tumors in the adjusted model (ER-; Supplementary Table 3) for asparagine (ER- tumors: OR (95%CI)=1.21 (0.99-1.47); ER+ tumors: OR (95%CI)=1.06 (0.94-1.18); p-interaction=0.03). Three additional metabolites were suggestively different by ER status: betaine (ER- tumors: OR (95%CI)=0.89 (0.73-1.09); ER+ tumors: OR (95%CI)=1.07 (0.96-1.20); p- interaction=0.02), 4-acetamidobutanate (ER- tumors: OR (95%CI)=0.89 (0.72-1.09); ER+ tumors: OR (95%CI)=0.97 (0.87-1.08); p-interaction=0.03), and histidine (ER- tumors: OR (95%CI)=1.06 (0.87-1.29); ER+ tumors: OR (95%CI)=1.01 (0.91-1.13); p-interaction=0.05).

In an exploratory analysis including metabolites with >10% missing values (N=19; 4 metabolites had >90% missingness), metoprolol (45% missing values) was nominally significantly associated with risk of breast cancer (likelihood-ratio test p-value=0.04; data not shown). Women with detectable metoprolol levels had a 23% higher risk (presence-absence indicator p-value=0.07) of breast cancer compared to women with undetectable metoprolol. However, among women with measured metoprolol, higher levels were associated with lower risk (OR per one unit increase in log-transformed and standardized metabolite levels =0.91, p-value=0.14). The remaining metabolites with high missingness were not associated with risk of breast cancer.

### Multivariable models of the joint association of all metabolites

The inverse association of piperine with risk of breast cancer met the threshold for statistical significance (p-value<0.05 and q-value<0.20) in all three simple (without adjustment for risk factors) multivariable models, Lasso, Elastic Net and Random Forests (Tables 2 and 3). In addition, DMGV and N2,N2-dimethylguanosine were detected in the Random Forests model with minimal adjustment, satisfying a q-value threshold of 0.05. Higher levels of C40:7 PE plasmalogen and creatine were associated with increased breast cancer risk in CLR Lasso models (q-value<0.20). These associations remained significant after further adjustment for risk factors (nominal p < 0.05) with the exception of N2,N2-dimethylguanosine in Random Forests (p=0.05) and piperine in Elastic Net (nominal p-value<0.05, q-value>0.2).

When all eleven identified metabolites were assessed together in an adjusted lasso CLR model, ten metabolites remained independently associated with risk of breast cancer. The direction of association in the lasso CLR model was consistent with the previous CLR and multivariable models except for N2,N2-dimethylguanosine who’s coefficient was estimated to be equal to zero, reflecting its high correlation with kynurenic acid and 2-aminohippuric acid (Figure 2).

**Table 2:**
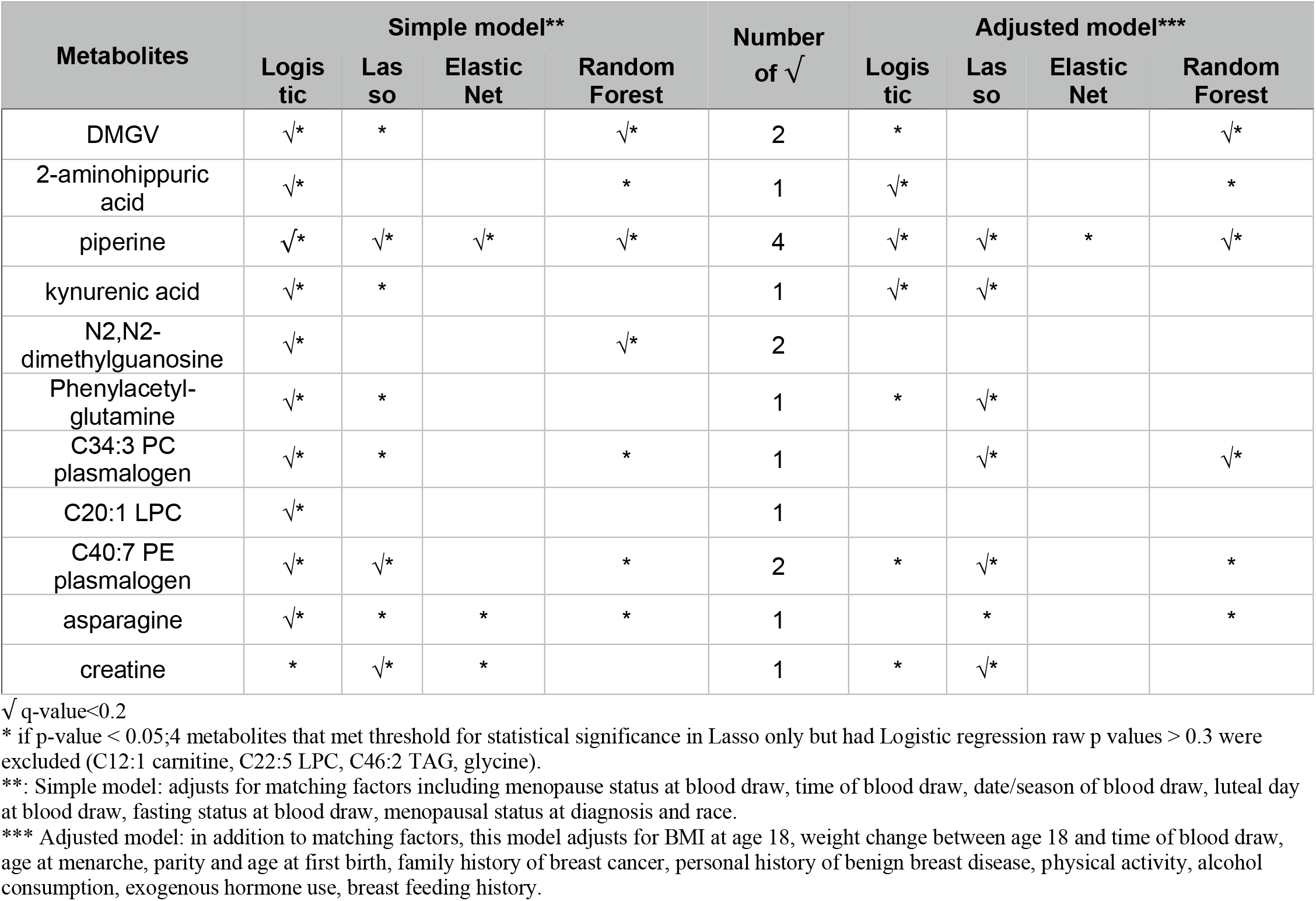
All metabolites that met q-value<0.2 in at least 1 primary model analysis that adjusts for matching factors.

**Figure 2:**
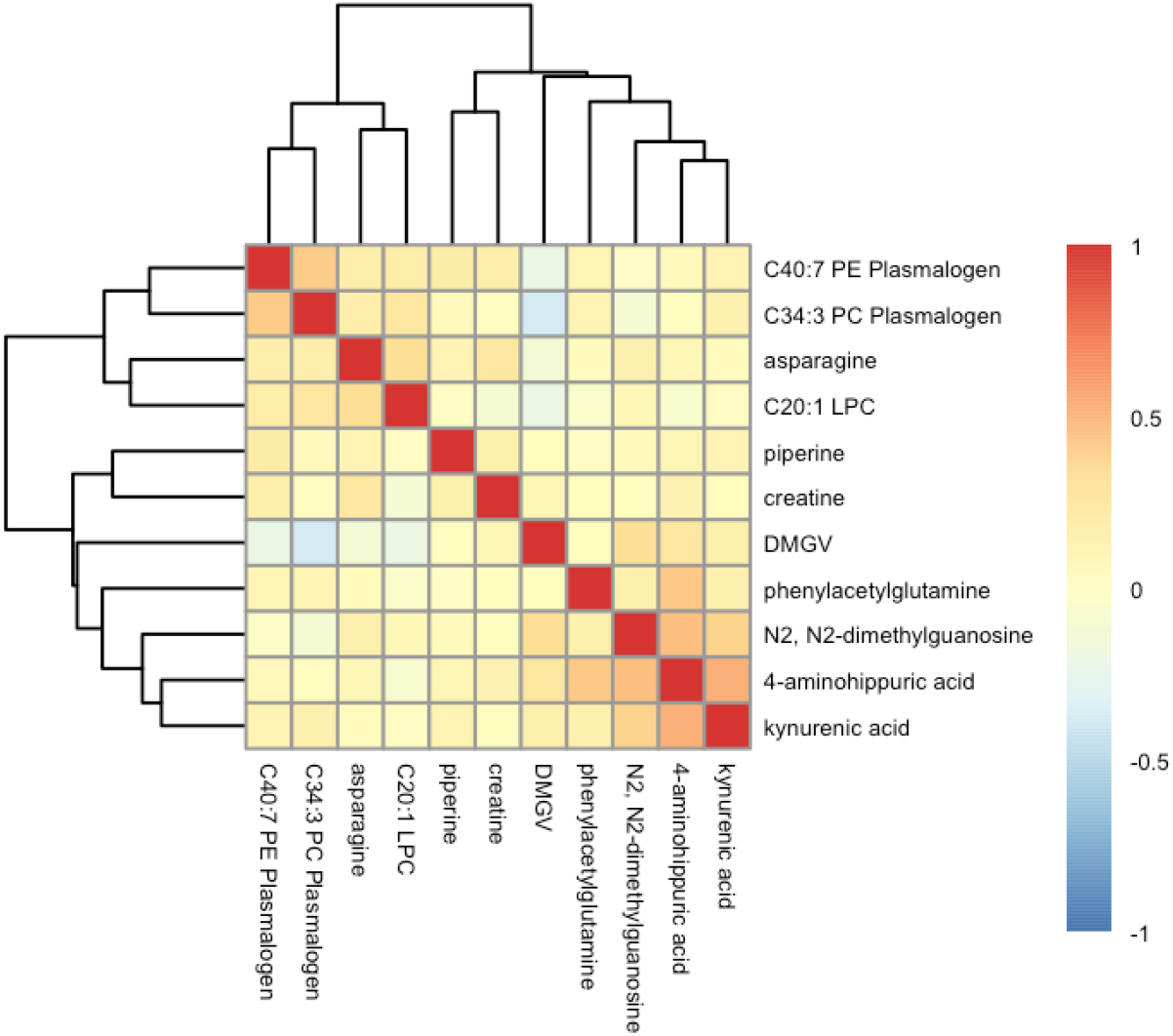
Heatmap of all pairwise correlations among the 11 metabolites associated with breast cancer risk. Positive correlations are shown in shades of red while inverse correlations are shown in shades of blue.

**Table 3:**
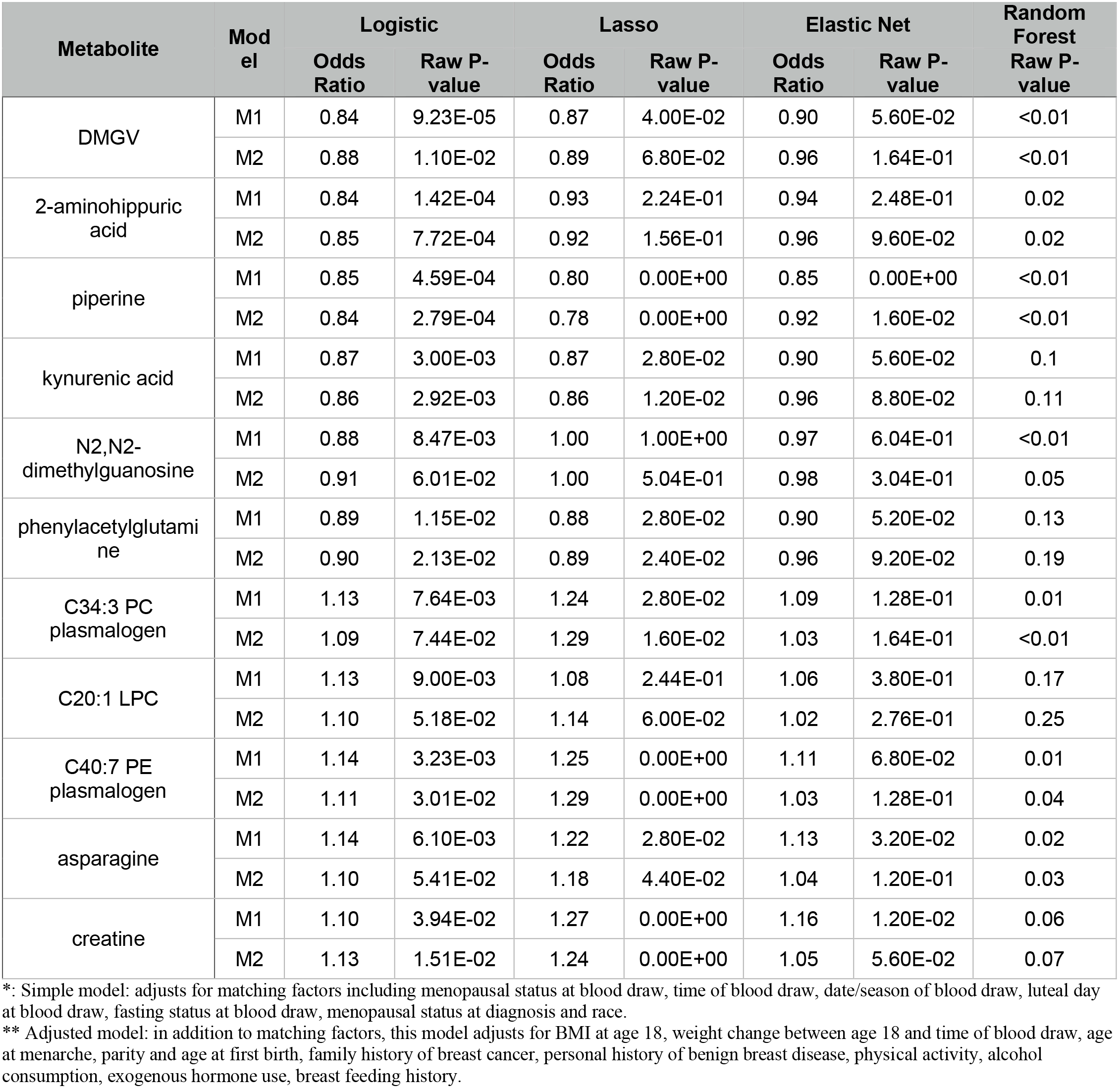
All metabolites that met q-value<0.2 in at least 1 primary model analysis that adjusts for matching factors. M1 is the simple model (accounting for matching factors) and M2 is the adjusted model (adjusting for matching factors and breast cancer risk factors).

## Discussion

We conducted a large-scale study of 207 circulating amino acid and amino acid-related metabolites, and risk of breast cancer in a nested case-control study (1057 cases and 1057 matched controls) within NHSII, a cohort of predominantly premenopausal women. Higher levels of six metabolites, 2-aminohippuric acid, DMGV, kynurenic acid, N2, N2-dimethylguanosine, phenylacetyl glutamine and piperine, were associated with lower breast cancer risk while higher levels of asparagine, creatine and 3 lipids, C20:1 LPC, C34:3 PC plasmalogen, C40:7 PE plasmalogen, were associated with increased breast cancer risk. Inverse associations between 2-aminohippuric acid, DMGV, kynurenic acid, phenylacetyl glutamine and piperine, and the positive association with C40:7 PE plasmalogen remained statistically significant after adjusting for established risk factors. Notably, associations between 2-aminohippuric acid, piperine and kynurenic acid remained significant even after multiple testing correction. Piperine, 2-aminohippuric acid and C40:7 PE plasmalogen were also selected in multivariable modeling approaches (Lasso, Elastic Net, and Random Forests). None of the metabolites showed heterogeneity by BMI, except DMGV. None of the metabolites showed heterogeneity by ER status, except asparagine.

Piperine is a polyphenol responsible for the pungency of black and long pepper and exhibits a wide range of properties: anti-diabetic, anti-inflammatory, immunomodulatory, reduction of insulin resistance, and enhanced drug bioavailability^33-35^. Piperine also inhibits tumorigenesis, tumor angiogenesis, cancer cell proliferation, cancer cell migration and invasion, and enhances apoptosis and autophagy^36^. Experimental and cell line studies identified anti-breast cancer specific mechanisms of action, including decreased matrix metalloproteinase 9 (MMP-9) and MMP-13 expression, induced apoptosis through activation of caspase-3 and inhibition of human epidermal growth factor receptor 2 (HER2) gene expression^37^. Synergetic effects of piperine and chemotherapy drugs (paclitaxel,doxorubicin), hormone therapy drugs (tamoxifen), radiotherapy, TRAIL- and nano-delivery-based therapy drugs (paclitaxel, rapamycine) were observed ^37^. Notably, piperine inhibited growth and motility^38^, and enhanced efficacy of TRAIL-based therapy^39^ in triple-negative breast cancer cells, the most aggressive breast cancer subtype. In a previous study of the associations of diet-related metabolites and breast cancer risk within the Prostate, Lung, Colorectal and Ovarian (PLCO) Cancer screening trial (n=1242, 621 cases), piperine was found to be modestly correlated with liquor consumption (correlation=0.16) and similar to our study, inversely associated with breast cancer risk (OR comparing 90^th^ versus 10^th^ percentile=0.74 (0.56-0.99,p=0.045)), after adjusting for BMI and other potential confounders^23^. Notably, PLCO women were postmenopausal at the time of blood collection suggesting that the association between piperine and breast cancer may be independent of menopausal status.

Dimethylguanadino valeric acid (DMGV), an organic keto acid, is the product of transamination of asymmetric dimethylarginine (ADMA), which inhibits nitric oxide signaling that is crucial to endothelial function—excess ADMA is associated with increased risk of cardiovascular disease^40,41^. Plasma DMGV is positively associated with incident coronary artery disease, cardiovascular mortality, nonalcoholic fatty acid liver disease, and type II diabetes^42,43^. Circulating DMGV is directly correlated with resistance to the metabolic benefits of exercise^44^. Physical activity, consumption of vegetables and red wine are associated with lower circulating DMGV, while sugar-sweetened beverage consumption is associated with higher circulating DMGV^43,45^. DMGV was associated with lower breast cancer risk in our study. The association between DMGV and breast cancer risk has not been previously assessed in prospective cohort studies. However, this metabolite was correlated with liver fat in the offspring cohort of the Framingham Heart Study (β=0.02, 95% CI: 0.018 - 0.022, p<10^-23^)^27^. In addition, in the same study, baseline DMGV levels were associated with higher risk of type 2 diabetes, with replication of this association in the Malmo Diet and Cancer study and the Jackson Heart Study^27^. Large prospective studies are required to validate the association between DMGV and breast cancer risk. If replicated, experimental studies will be needed to understand the complex relationship between DMGV, diet, physical activity, CVD, type II diabetes, and breast cancer. Of note is that this analysis was in predominantly premenopausal women, among whom adiposity is also inversely associated with risk of breast cancer for reasons that are still not fully understood^46^.

Although the measurement platform used here was optimized to measure amino acids and related metabolites, not lipids, our analysis included a small number of lipids and identified a few significant associations. Plasmalogens are a subclass of phospholipids (components of the cell membrane and involved in cell signaling and cell cycle regulation^47^) that constitute 15-20% of all phospholipids in cell membranes^48^. Plasmalogens have head groups that are usually either phosphatidylcholine (PC plasmalogens) or ethanolamine (PE plasmalogens), and are characterized by an ether bond to an alkenyl group in the sn-1 position while the sn-2 position is usually occupied by polyunsaturated fatty acids^48^. Certain cancers exhibit altered plasmalogen levels: circulating plasmalogens are depressed in pancreatic cancer patients^49^ and increased in gastric carcinoma patients^50^ compared to healthy controls. Our study identified two plasmalogens, C34:3 PC plasmalogen and C40:7 PE plasmalogen, associated with increased risk of breast cancer. The fatty acid component of specific lipids may also reflect dietary or metabolic processes; notably C40:7 PE plasmalogen is highly unsaturated but the position of the double bonds cannot be determined in this metabolomics assay. Among 74 women with breast cancer, the levels of the majority of measured phospholipids (LPC, LPE, PC and PE) were higher in tumor tissue when compared to normal breast tissue samples^47^. Similar trends were observed in another study comparing tumor to normal tissue in 257 participants with breast cancer, and these lipids were correlated with cancer progression and patient survival^51^. Contrary to our findings, in the European Prospective Investigation into Cancer (EPIC) cohort, PC plasmalogens, including C34:3 PC plasmalogen, were inversely associated with risk of breast cancer^18,19^. However, neither study stratified their results by menopausal status, thus making a direct comparison difficult. Additional studies are needed to evaluate how plasmalogens are associated with risk of breast cancer and if this relationship is modulated by menopausal status.

LPCs are derived from phosphatidylcholines after hydrolysis of one of the fatty acid groups. In the liver, LPCs upregulate genes involved in cholesterol biosynthesis, while circulating LPCs activate many inflammatory and oxidative stress signaling pathways, and are associated with inflammatory diseases such as atherosclerosis and multiple sclerosis^52^. Circulating LPCs have mixed associations with certain cancers. For instance, circulating LPCs are elevated in ovarian cancer patients but depressed in leukemia patients relative to healthy controls^53^; LPCs showed inverse associations with risk of endometrioid and clear cell ovarian tumors, with stronger inverse associations among premenopausal women, in NHS and NHSII^15,16^. While most LPCs measured in EPIC were inversely associated with risk, one of our top hits was LPC C20:1 that was positively associated with risk^18^. In an earlier study nested within the EPIC cohort of 774 participants including 362 breast cancer cases, C18:0 LPC was identified as inversely associated with breast cancer risk, after adjusting for potential risk factors ^19^, though analyses were not stratified by menopausal status.

Our study identified three amino acid derivatives associated with breast cancer risk. High levels of phenylacetylglutamine were associated with decreased breast cancer risk while high levels of asparagine and creatine were associated with increased risk. Phenylacetylglutamine is formed from phenylacetate and glutamine and is found as a normal constituent of human urine^54^. Phenylacetylglutamine is a host microbiome cometabolite associated with bacterial phenylalanine metabolism^55-58^. *Clostridium difficile, F. prausnitzii, Bifidobacterium, Subdoligranulum*, and *Lactobacillus* are all positively associated with hippuric acid^57,59^, while *Bifidobacterium* is positively associated with phenylacetylglutamine and microbes of the *Christensellaceae, Ruminococcaceae*, and *Lachnospiracaea* families are negatively associated with phenylacetylglutamine^56,60^. While not directly linked to breast cancer risk, high serum levels of phenylacetylglutamine is a potential early marker of kidney dysfunction in chronic kidney disease^60^. Glutamine, a precursor to phenylacetylglutamine, has been associated with breast cancer risk in a nested case-control study within the French Su.Vi.Max cohort (n=211 cases). High levels of glutamine were associated with increased risk (OR per SD increase =1.33, 95% CI: 1.07-1.66) and this association persisted among the subgroup of premenopausal women (p for interaction = 0.003)^20^. In a nested case-control study within the EPIC cohort (n=1624 cases), asparagine was inversely associated with breast cancer risk (OR=0.87 per SD increase, 95% CI: 0.80-0.95, FDR p=0.06), in contrast to the direction of association in our study^18^. However, the EPIC study participants were overwhelmingly (> 70%) postmenopausal at the time of blood collection, in contrast to our population with 80% premenopausal women. Differences in the menopausal status may partially explain the observed opposite directions of association between asparagine and breast cancer risk.

Creatine is obtained from meat consumption and synthesized endogenously from arginine, glycine, and methionine. Most creatine is found in skeletal muscle, and a significant amount is also found in the brain. Omnivores obtain roughly 50% of their daily creatine from meat and 50% is biosynthesized, while vegetarians biosynthesize most of their creatine^61^ and have significantly lower muscular creatine levels than meat eaters^62^. Creatine is broken down to creatinine in a first-order reaction, the rate of which decreases with age and decreased muscle mass^63^. Creatine/creatinine metabolism plays an important role in energy metabolism in skeletal muscle tissue, and thus disturbances in this pathway are associated with many muscle diseases, whether as a cause or consequence^64^. To the best of our knowledge, no previous work has reported a link between creatine and breast cancer risk. However, the association we observed in this analysis is consistent with the positive association between red meat and risk of breast cancer among premenopausal women in the NHSII cohort^65^.

Kynurenic acid and 2-aminohippuric acid are benzenoids inversely associated with breast cancer risk in our study. 2-amminohippuric acid is a glycine conjugate of anthranilic acid and can be synthesized in the liver^66^, but little is known about its biological function. Both kynurenic acid and 2-aminohippuric acid are part of the kynurenine branch of the tryptophan pathway, an essential amino acid and a precursor to many biologically active metabolites^67,68^. Studies of the function of kynurenic acid suggest pleiotropic roles in disease. On the one hand, kynurenic acid has been shown to have anti-inflammatory and anti-ulcerative properties in animal models, as well as antioxidative properties *in vitro* in human cells^69^. On the other hand, circulating kynurenic acid was associated with increased risk of insulin resistance^70^. Kynurenic acid acts as both an anti-inflammatory and immunosuppressive factor which in turn allows tumor proliferation^71^. Tryptophan metabolism plays an important role in tumor progression and malignancy with several cancers expressing tryptophan-degrading enzymes such as IDO1^72,73^. However, the direct role of kynurenic acid remains unclear, with both proliferative and antiproliferative effects on human glioblastoma cells, an antiproliferative effect on human colon adenocarcinoma cells, and decreased DNA synthesis and inhibited migration in both cell types^69^. Neither kynurenic acid nor 2-aminohippuric acid have been previously associated with breast cancer risk.

N2,N2-dimethylguanosine (DMGU) is a purine nucleoside and a primary degradation product of transport RNA. Elevated circulating DMGU levels may indicate cellular stress and is associated with several diseases, including pulmonary arterial hypertension^74^, solid tumors^75^, incident type II diabetes^76^, and all-cause mortality^77^. Elevated levels of N2, N2-dimethylguanosine were found in patients with acute leukemia and breast cancer^78^. Elevated circulating levels of this metabolite are associated with lower risk of breast cancer in our study.

Our study has several strengths and limitations. Notably, we conducted a prospective analysis of amino acid and amino acid-related metabolomics and risk of breast cancer among a large number of predominantly premenopausal women. We had detailed information on sample collection characteristics and risk factors which we included in our statistical approaches. Although metabolomics was measured at only one point in time, the identified metabolites are reasonably stable over time^29^ (ICCs or correlation over 1-2 years ≥0.75 for 9 metabolites; no data are available for 2-aminohippuric acid and C20:1 LPC). Our cohort consisted of registered nurses, a group that are not representative of the general population (e.g. social economic status), however there is no evidence suggesting that breast carcinogenesis is different in this group of women. While we had reasonable power in most of our analyses, we had limited power among ER negative tumors. Lastly, the uniqueness of our data measured among predominantly premenopausal women make replication studies challenging. We conducted this analysis in a hypothesis generating framework and hope that other cohorts will follow and analyze metabolomics data stratifying by menopausal status.

In summary, we identified several metabolites associated with risk of breast cancer among premenopausal women. Increased circulating levels of piperine, 2-aminohippuric acid and kynurenic acid are associated with lower risk of breast cancer, independent of established risk factors and after accounting for testing multiple hypotheses. Additional prospective cohort studies are needed to assess these associations considering menopausal status. If these findings are validated, experimental studies are warranted to understand the underlying biological mechanisms driving changes in metabolite levels.

## Data Availability

Data will be made available upon reasonable request.

## Acknowledgements

This study was funded by the National Cancer Institute (R01 CA050385, UM1 CA186107, P01 CA087969, R01 CA49449, U01 CA176726, R01 CA67262). We would like to thank the participants and staff of the Nurses’ Health Studies for their valuable contributions as well as the following state cancer registries for their help: AL, AZ, AR, CA, CO, CT, DE, FL, GA, ID, IL, IN, IA, KY, LA, ME, MD, MA, MI, NE, NH, NJ, NY, NC, ND, OH, OK, OR, PA, RI, SC, TN, TX, VA, WA, WY. The authors assume full responsibility for analyses and interpretation of these data.

## Notes

### Competing Interest Statement

The authors have declared no competing interest.

### Clinical Trial

This is not a clinical trail.

### Author Declarations

The study protocol was approved by the institutional review boards of the Brigham and Women's Hospital and Harvard T.H. Chan School of Public Health, and those of participating registries as required.

